# Trabectedin in the Treatment of Soft Tissue Sarcoma: Real-world Data on Effectiveness, Safety, and Financial Implications from a European Comprehensive Cancer Centre

**DOI:** 10.1101/2025.08.22.25334220

**Authors:** Jean-Stéphane Giraud, Sarah Watson, Alexandre Acramel, Valérie Laurence, Dimitri Tzanis, Sylvie Bonvalot, Sophie El Zein, Nayla Nicolas, Cyrille Cros, Romain Desmaris, Clément Bonnet

## Abstract

**Background:** Soft tissue sarcomas (STS) comprise over 150 histological subtypes, with advanced cases showing poor prognosis (5-year survival <10%). Trabectedin, a synthetic alkaloid, is frequently used after anthracycline-based chemotherapy failure. Despite the withdrawal of reimbursement in France in 2018 due to debated efficacy and safety, it remains in clinical use, imposing financial strain on hospitals.

**Methods:** This retrospective single-center study evaluated trabectedin’s efficacy, safety, and cost in 68 patients treated between 2019 and 2023.

**Results:** L-sarcomas accounted for 78% of cases, including uterine leiomyosarcomas (n=16), soft-tissue leiomyosarcomas (n=17), and myxoid liposarcomas (n=8). Non-L-sarcomas (22%) included mostly synovial sarcomas. The overall disease control rate was 71%, with a median progression-free survival (PFS) of 4.1 months. Subtype-specific median PFS was 6.8 months for liposarcomas (11.3 for myxoid vs. 4.5 for other subtypes), 3.1 months for leiomyosarcomas (3.4 months for uterine vs 3.1 for soft-tissue), and 2.4 months for non-L-sarcomas. Patients received a median of 5 cycles (range: 1–38), with an average total dose of 16 mg [2 – 81], and an average hospital cost of €9,900. Adverse events occurred in 91%, mainly hematological; cardiac toxicity was seen in 9%.

**Conclusion:** Despite limited reimbursement, trabectedin remains a relevant treatment, particularly in L-sarcoma management.

## Background

Soft tissue sarcomas (STS) represent a heterogeneous group of tumors comprising over 150 histological subtypes (1,2). They are considered rare, with an overall incidence rate ranging from 4 to 5 cases per 100,000 individuals per year, while many histotypes occur at a rate as low as 0.1 cases per 100,000 (3,4). Patients diagnosed with locally advanced or metastatic STS have a less than 10% chance of overall survival (OS) at five years (5). The most common subtypes in adults, i.e. liposarcomas and leiomyosarcomas, are referred to as L-sarcomas. Non-L-sarcomas encompass a diverse array of other STS subtypes, including translocation-related sarcomas (6).

The standard systemic treatment for advanced STS typically involves first-line chemotherapy that includes an anthracycline such as doxorubicin, sometimes combined to the alkylating agent ifosfamide (7). The 2021 clinical practice guidelines from the European Society for Medical Oncology and the European Reference Network for rare adult solid cancers recommend trabectedin, a synthetic marine-derived antitumor alkaloid, as a second-line treatment option for advanced STS (8).

Trabectedin (Yondelis^®^, *PharmaMar*) was approved in 2017 by the European Medicines Agency for patients with advanced STS who had failed anthracyclines and ifosfamide or were not suitable to receive such agents (9). This approval was based on a randomized phase III study (ET743-SAR-3007, NCT01343277) that compared the safety and efficacy of trabectedin 1.5 mg/m^2^ as a 24-hour continuous infusion once every three weeks to dacarbazine (1,000 mg/m^2^) administered every three weeks mainly on liposarcoma and leiomyosarcoma patients (10). In this study, no difference in overall survival (primary endpoint) was evidenced: 13.7 months vs. 13.1 months (Hazard Ratio = 0.927, Confidence Interval 95% [0.748-1.150], p=0.49) (10). The process for reimbursing oncology drugs in Europe is complex and varies among Member States (11). In 2018, the French National Authority for Health ended the definitive reimbursement for this indication (12). The main reasons for this decision included: (i) a lack of benefit in OS compared to dacarbazine in liposarcoma and leiomyosarcoma (10); (ii) a lack of benefit in OS compared to supportive care in various histological subtypes including liposarcoma and leiomyosarcoma (13); and (iii) concerns regarding the drug’s safety profile. Furthermore, trabectedin’s place in the therapeutic arsenal has not been fully defined compared to other comparators such as pazopanib (14) and eribulin (15).

Despite these challenges, trabectedin continues to be utilized in clinical practice with a consequent non-reimbursed cost, therefore impacting hospital budgets (16). We aimed to update existing cohort studies (Supplemental Table 1) (10,13,17–28) by assessing trabectedin efficacy, safety, and costs in a specialized referral single-center cohort of “real-life’’ patients with STS.

**Table 1.** Baseline characteristics of the population.

| Variable | Value |
| --- | --- |
| Age, years old: median [minimum – maximum] | 47 [12-74] |
| Gender, n (%) |  |
| Male | 30 (44) |
| Female | 38 (56) |
| BMI, kg.m<sup>-2</sup>: mean (SD) or n (%) | 25.7 (5.9) |
| <18 | 4 (6) |
| 18-24 | 30 (44) |
| 25-29 | 19 (28) |
| ≥ 30 | 13 (19) |
| Unknown | 2 (3) |
| ECOG Performance Status, n (%) |  |
| ≥2 | 10 (15) |
| 0-1 | 56 (82) |
| Unknown | 2 (3) |
| Sarcoma subtypes, n (%) |  |
| Extraskeletal myxoid chondrosarcoma | 1 (1.5) |
| Desmoplastic small round cell tumors | 2 (3) |
| Sclerosing epithelioid fibrosarcoma | 1 (1.5) |
| Leiomyosarcoma | 33 (49) |
| Liposarcoma | 20 (29) |
| Other | 2 (3) |
| Synovialsarcoma | 9 (13) |
| Leiomyosarcoma subtypes, n (%) |  |
| Soft tissue | 17 (52) |
| Uterine | 16 (48) |
| Liposarcoma subtypes, n (%) |  |
| Dedifferentiated | 7 (32) |
| Myxoid | 10 (45) |
| Pleomorphic | 1 (5) |
| Well-differentiated | 4 (18) |
| Metastasis sites, n (%) |  |
| Bone | 15 (22) |
| Brain | 1 (1) |
| Cutaneous/subcutaneous | 5 (7) |
| Lymph nodes | 12 (18) |
| Liver | 24 (35) |
| Muscular | 6 (9) |
| Other | 6 (9) |
| Pancreatic | 5 (7) |
| Pelvic | 2 (3) |
| Peritoneal | 14 (21) |
| Lung | 45 (66) |
| Baseline biological data: mean (SD) |  |
| Hemoglobin (g/dL) | 12.3 (1.7) |
| Neutrophils (G/L) | 5.1 (4.3) |
| Lymphocytes (G/L) | 1.3 (0.6) |
| Albumin (g/L) | 38.8 (5.5) |

SD: standard deviation.

### Patients and Methods

We conducted a retrospective non-interventional study at Institut Curie, a European comprehensive cancer center member of the European Reference Network for sarcoma.

#### Patient population

Medical records of patients with histologically proven STS who had received at least one cycle of trabectedin between 2019 and 2023 were reviewed. All STS subtypes were included. The study was performed in accordance with the Declaration of Helsinki. Eligible patients had signed a non-opposition consent form.

#### Data collection

Clinical and biological data were collected from the patient’s electronic record system and the CHIMIO^®^ chemotherapy prescribing software (Computer Engineering, Paris, France).

Clinical data were collected at baseline, including age, gender, Eastern Cooperative Oncology Group (ECOG) Performance status, pathological subtypes, and metastatic status with metastatic sites. The collected biological data included hemoglobin, neutrophils, lymphocytes, and albumin.

#### Trabectedin use

Prior treatment history, number of cycles administered and applied doses, dose reductions, the date and the reasons for treatment discontinuation were collected. Patients were regularly assessed clinically and radiologically using the Response Evaluation Criteria in Solid Tumors (RECIST 1.1). Toxicities were graded using the National Cancer Institute – Common Terminology Criteria for Adverse Events version 4.0 (CTCAE). The primary endpoint was Progression-free survival (PFS), defined as the time from the first trabectedin injection to clinical and/or radiological disease progression. Patients were classified as having disease control if they had a complete response (CR), partial response (PR), or stable disease (SD). The disease control rate (DCR) was defined as the sum of CR, PR, and SD at the best response. The objective response rate (ORR) was defined as the sum of CR and PR.

#### Economic analysis

The prices of trabectedin vials were obtained from the public procurement agreements of our central purchasing body, supported by the UNICANCER federation. We valued the “diagnosis-related groups” using a national survey on hospital costs with a valuation year set as 2022 (29).

### Statistical analysis

Descriptive statistics were performed using mean [minimum – maximum] or mean (standard deviation) for quantitative variables, and numbers and percentages for qualitative variables. PFS were estimated with the Kaplan-Meier method, and the log-rank test was used to compare survival curves between subgroups. The difference in PFS between the sarcoma subtypes was compared the hazard ratio (HR), with its 95% confidence interval (CI) calculated from a Cox regression model with a single covariate. All tests were two-tailed, and the significance level was set at p < 0.05. Statistical analyses were performed with R statistical software (version 4.2.3).

## Results

### Patient characteristics

We included 68 patients: 38 women and 30 men, with a median age of 47 years [12 – 72] at the time of trabectedin initiation. Most STS subtypes were L-sarcomas (53/68, 78 %), primarily leiomyosarcomas (33/53, 62 %) – 16 uterine leiomyosarcomas and 17 soft-tissue leiomyosarcomas - followed by liposarcomas (20/53, 38 %). Among patients with liposarcomas, the histological subtypes were myxoid (8/20, 40 %), dedifferentiated (7/20, 35 %), well-differentiated (4/20, 20 %) and pleomorphic (1/20, 5 %). Fifteen patients (22 %) had non-L-sarcomas, mostly synovial sarcomas (9/15, 60 %) (**Table 1**). All patients presented metastatic disease at the time of trabectedin initiation, with 21 patients (31 %) being metastatic at diagnosis. The most common metastatic sites were lungs (45/68, 66 %), liver (24/68, 35 %), and bone (15/68, 22 %). The locations of other metastases are shown in **Table 1**. The majority of patients (56/68, 82 %) had an ECOG performance status of 0 or 1.

### Treatment characteristics

Trabectedin was mainly used in a second-line treatment (30/68, 44%) (Range: 1-7) (**Figure 1A**). Prior to trabectedin, 60 patients (86 %) had received doxorubicin-based chemotherapy. Four patients were treated with an association of trabectedin and doxorubicin. Patients received a mean of 8 cycles [1 – 38] (median: 5) of trabectedin with the majority receiving 2 cures (18/68, 26 %) (**Figure 1B**). Thirteen (19%) patients received more than 10 cycles of trabectedin, mainly with myxoid liposarcomas (5/13, 38 %). Twelve patients (17%) were still receiving trabectedin at the time of data censoring. Five patients (7%) underwent therapeutic rechallenge with trabectedin.

**Figure 1.**
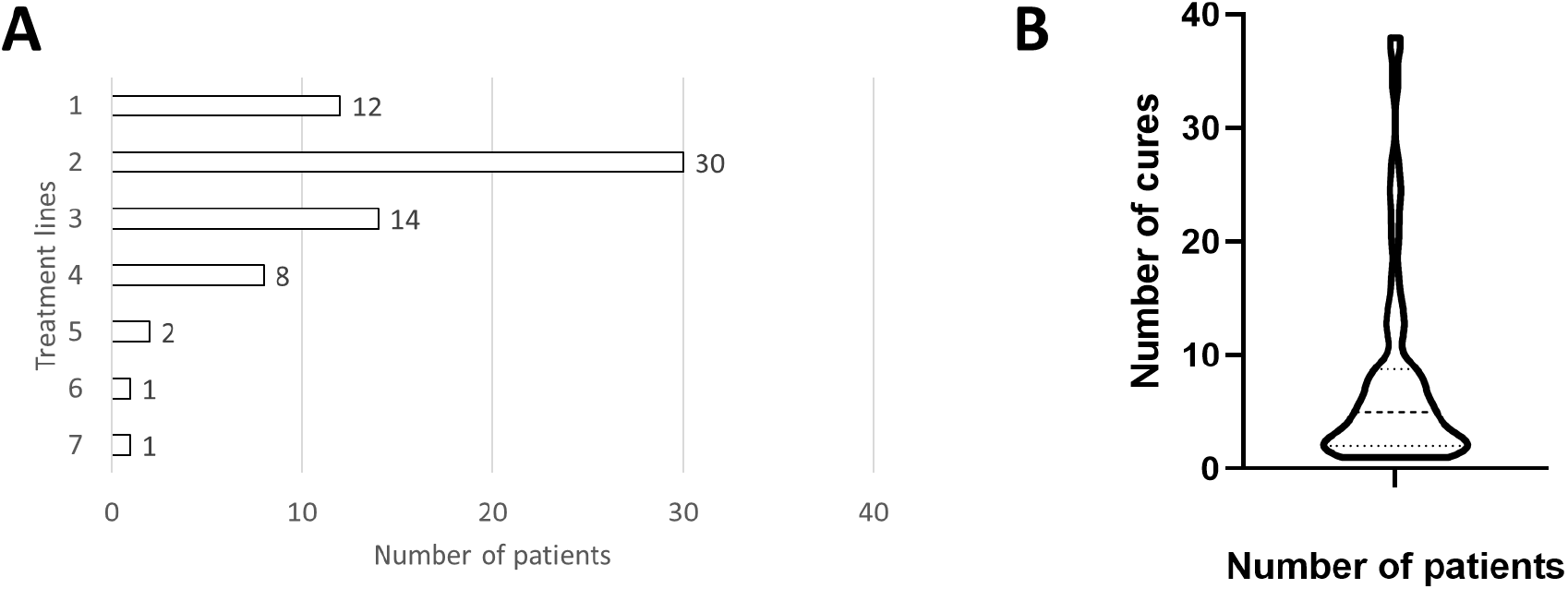
Trabectedin use: lines of treatment and number of cycles

### Costs of treatment

The price per milligram of trabectedin vials decreased drastically between 2019 and 2023, from €770 to €180 excluding tax (**Supplementary Figure S1**). In this cohort, the average total dose administered was 16 mg [2 – 81], resulting in an average cost of €9,900 [342-62,288] per patient for the hospital. During the period studied, the mean cost per trabectedin administration was €1,159 [77-2,887].

### Response assessment

In all subtypes combined, the DCR was 71 % and the ORR was 26 %, with 2 CR (3 %), 16 PR (24 %), and 30 SD (44 %) (**Supplementary Figure S2, Supplementary Table S1**). Seventeen patients (24 %) showed tumor progression as best reponse when receiving trabectedin, including 30 % (n=5) of non-L-sarcoma patients (**Figure 2**). The DCR was 70% in leiomyosarcomas (69% for uterine, 71% for soft-tissue), 80% in liposarcomas (88% for myxoid and 75% for other subtypes), and 60% in non-L-sarcomas. The ORR was 27% in leiomyosarcomas (31% for uterine, 24% for soft-tissue), 25% in liposarcomas (50% for myxoid and 8% for other subtypes), and 27% in non-L-sarcomas (**Table 2**). Causes of treatment discontinuation were disease progression (46/56, 82 %) and tolerability issues (6/56, 11 %). The main therapeutic options implemented after trabectedin discontinuation were gemcitabine (10/56, 18 %), cyclophosphamide (9/56, 16 %), palliative care (7/56, 13 %), and pazopanib (6/56, 11 %). For non-L-sarcomas, the subsequent anticancer treatments included pazopanib (3/15, 20 %) and oral cyclophosphamide (3/15, 20 %).

**Figure 2.**
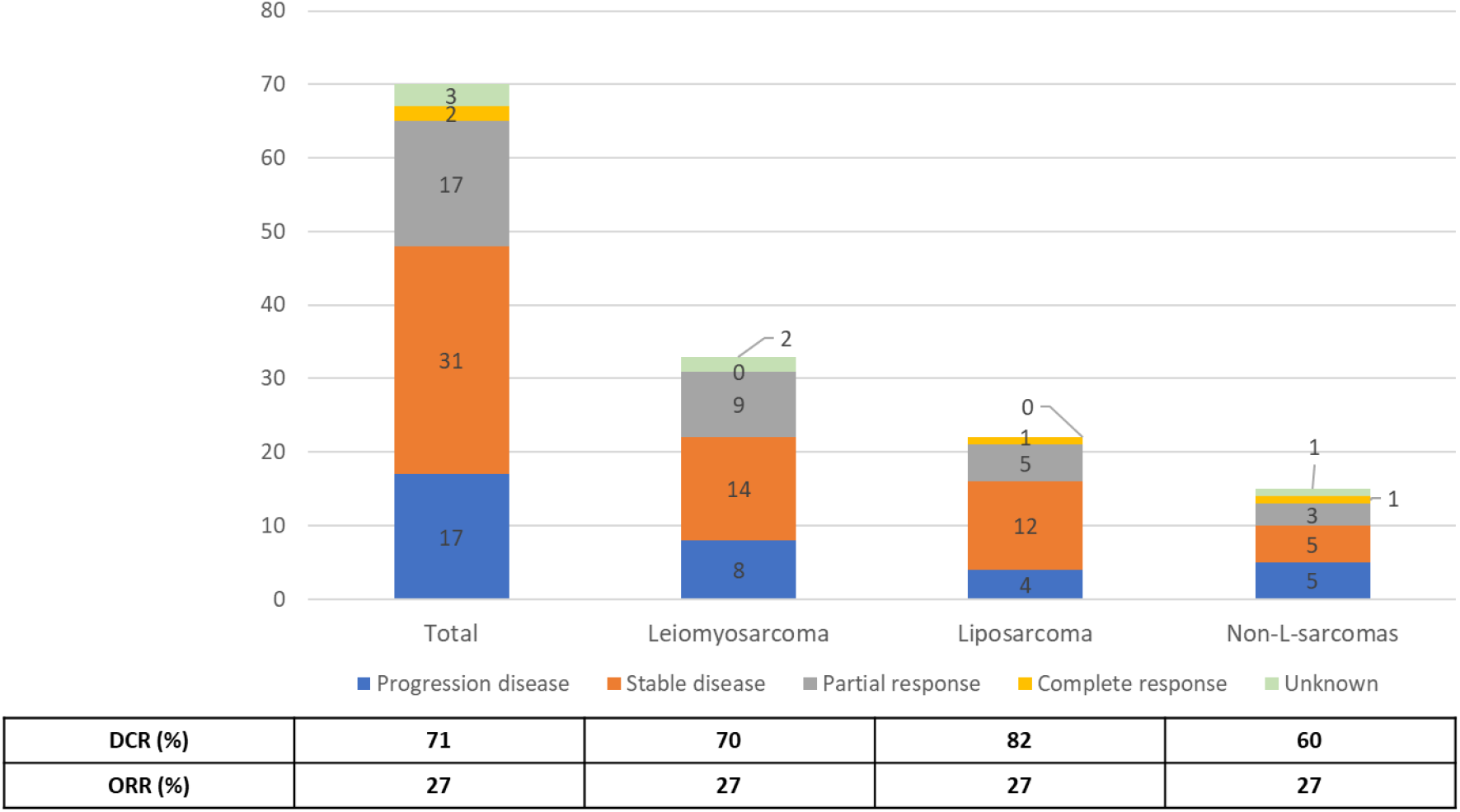
Disease-control and overall response rates due to trabectedin according to STS subtypes. DCR: Disease-control rates. ORR: Overall response rates.

**Table 2.** Trabectedin best response assessment in L-sarcomas.

| N (%) | Leiomyosarcoma (n=33) |  | Liposarcoma (n=20) |  |
| --- | --- | --- | --- | --- |
| Subtypes | Uterine (n=16) | Soft-tissue (n=17) | Myxoid (n=8) | Other (n=12) |
| Complete response | 0 (0) | 0 (0) | 1 (13) | 0 (0) |
| Partial response | 5 (31) | 4 (24) | 3 (37) | 1 (8) |
| Stable disease | 6 (38) | 8 (47) | 3 (37) | 8 (67) |
| Progressive disease | 4 (25) | 4 (24) | 1 (13) | 3 (25) |
| Unknown | 1 (6) | 1 (5) | 0 (0) | 0 (0) |
| Objective response rate | 5 (31) | 4 (24) | 4 (50) | 1 (8) |
| Disease control rate | 11 (69) | 12 (71) | 7 (88) | 9 (75) |

### Survival data with subgroup analysis

The median PFS was 123 days (SD: 278), equivalent to 4.1 months. The median PFS for patients with leiomyosarcoma, liposarcoma, and non-L-sarcoma was, 3.1, 6.8, and 2.4 months respectively (**Table 3**). Patients with uterine or soft-tissue sarcomas have the same median PFS (3.4 vs 3.1 months). Although patients with myxoid liposarcomas appear to have better PFS (11.3 months) than other subtypes of liposarcomas (4.5 months), the difference is not significant (**Supplementary Table S2**). No significant difference (p>0.05) in PFS was observed when comparing (i) liposarcoma, leiomyosarcoma, and non-L-sarcoma (**Supplementary Figure S3A**), (ii) myxoid liposarcomas and other liposarcoma subtypes (**Supplementary Figure S3B**), and (iii) uterine leiomyosarcomas and soft-tissue leiomyosarcomas (**Supplementary Figure S3C**).

**Table 3.** Compared survival between Leiomyosarcoma and Liposarcoma or Non-L-sarcoma.

|  | Number of patients | PFS: mean (SD), days | PFS: median, months | HR [95%CI] | p-value |
| --- | --- | --- | --- | --- | --- |
| Leiomyosarcoma | 33 | 154 (160) | 3.1 | / | / |
| Liposarcoma | 22 | 393 (348) | 6.8 | 0.558 [0.287, 1.087] | 0.0863 |
| Non-L-sarcoma | 15 | 128 (127) | 2.4 | 1.155 [0.562, 2.373] | 0.6946 |

HR: hazard ratio; CI: confidence interval

### Safety

Toxicities were reported in 62 patients (91%). The most common toxicities were hematological, asthenia, nausea, constipation, and hepatic cytolysis (**Figure 3**). The most frequent grade 3–4 toxicities observed were hematological, mainly neutropenia (14/19, 74%).

**Figure 3.**
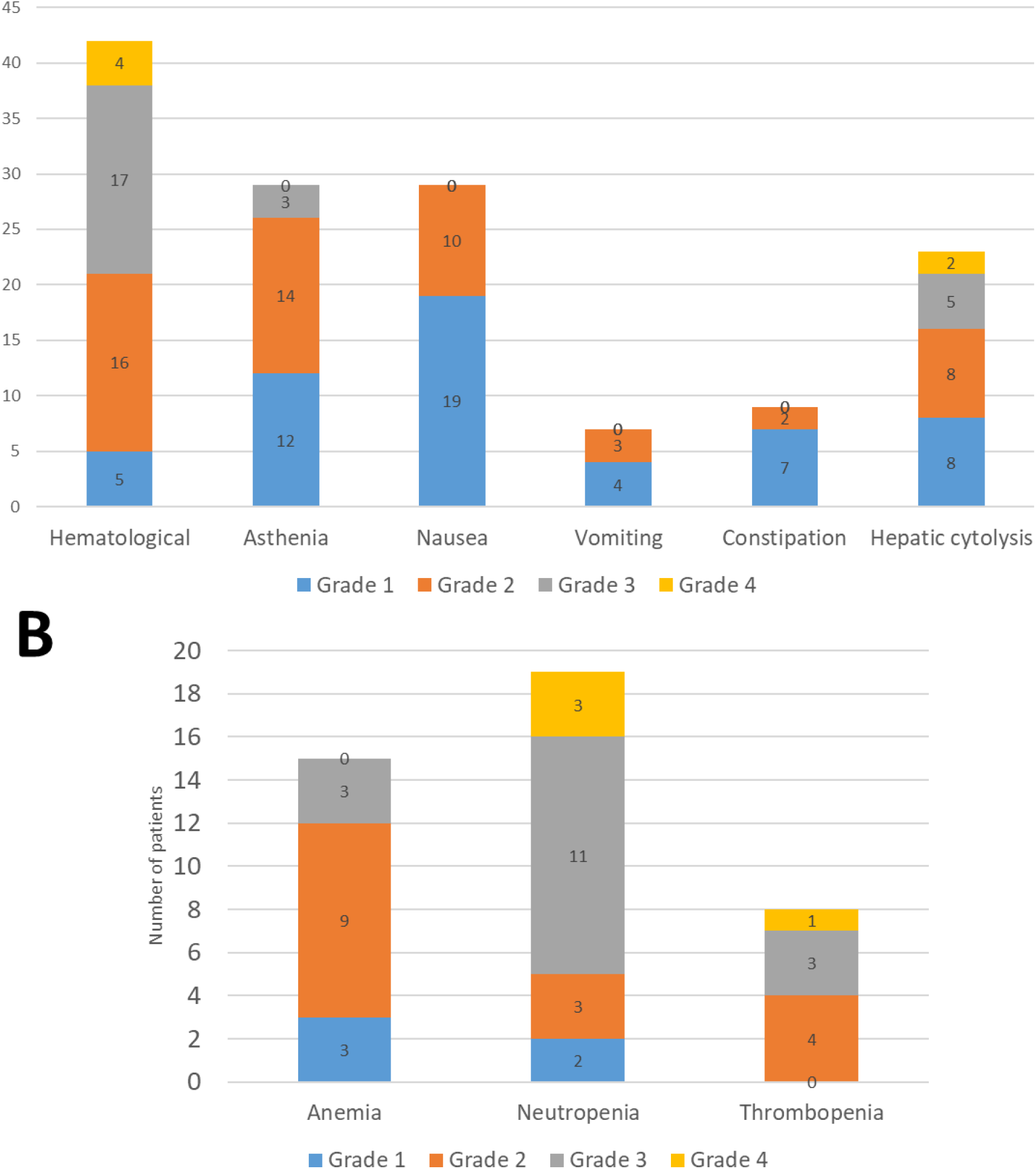
Toxicities reported with trabectedin use

In total, 22 patients (32%) had their trabectedin dose reduced (down to 0.8mg/m^2^). Additionally, 26 patients (38%) had their courses of treatment cycles spaced out, typically every four weeks.

Six patients experienced cardiac toxicity, including two cases of acute heart failure and one case of septic and/or cardiogenic shock. Only one of these six patients received co-administration of doxorubicin with trabectedin.

A patient experienced trabectedin extravasation, trabectedin being a chemotherapy drug known to be a vesicant, with an estimated extravasated volume of approximately 100mL. Initially, the patient reported paresthesias. The infusion was stopped immediately, and aspiration along with the removal of the Hubert needle was performed. A few hours later, the patient reported edema of the right shoulder, localized redness, vesicles without skin necrosis, but no immediate pain. The erythematous area was delimited, and a cold pack was applied. In the operating room, the procedure included subcutaneous aspiration, abundant saline irrigation, and unsutured incisions to facilitate the evacuation of any remaining fluid. The vesicles resolved within a few days. Although healing improved slowly with daily home dressings, the patient continued to experience extreme local pain for an extended period, requiring analgesic treatment such as oral morphine.

## Discussion

This retrospective study updates previously published cohort studies on the use of trabectedin in STS (Supplementary Table 1). Trabectedin is effective mainly in L-sarcoma, and myxoid liposarcoma (n=8) was the histology subtype that benefited most from it (median PFS: 11.3 months, disease-control rate: 88%). Here, 22% (n=15) of the cohort involved non-L-sarcomas, which is interesting given the rarity of these cancers. Our findings show a DCR of 60% and a median PFS of 2.4 months for non-L-sarcomas. However, in all subtypes, we caution regarding cardiac risks associated with trabectedin, which are currently under-reported and warrant increased cardiologic monitoring.

In this study, trabectedin was used mainly as a second-line treatment in accordance with the marketing authorization after doxorubicin, as recommended by the 2021 clinical practice guidelines from the European Society for Medical Oncology and the European Reference Network for rare adult solid cancers (8). The median PFS for all histological subtypes was 4.1 months, which is a little higher than what is typically reported in the literature, around 3 months (**Supplementary Table S3**). A reference study is the T-SAR study (NCT02672527), a phase III, open-label, randomized, multicenter trial that evaluated the efficacy and safety of trabectedine (n=52) *versus* supportive care (n=51) in adult patients with advanced STS resistant or refractory to anthracyclines and ifosfamide. The median PFS (primary endpoint) was 3.1 months (IC95% [1.80 - 5.85]) in the trabectedin group *versus* 1.5 months (IC95% [0.92 - 2.63]) in the supportive care group (HR=0.39, IC95% [0.24-0.64]) (13). In our cohort, L-sarcomas exhibited the best response to trabectedin treatment, consistent with the literature (Supplementary Table I). Patients with myxoid liposarcomas have benefited the most from trabectedin: majority sub-type where patients have had more than 10 trabectedin cycles, highest median PFS, and highest DCR. Myxoid liposarcomas are indeed known to be particularly chemosensitive to trabectedin (30). Trabectedin mostly provides disease stabilization, with a relatively low objective response rate (27%, consistent with the literature). Trabectedin treatment is also feasible for non-L-sarcomas, the median PFS was 2.4 months higher than the 1.8 months observed in the T-SAR study, but DCR was consistent (60% *versus* 69%) (13).

The primary toxicities seen in our cohort align with the most common trabectedin-related adverse events identified in a literature review (31): nausea, fatigue, vomiting, reversible myelosuppression (neutropenia, thrombocytopenia), and transient reversible elevations in transaminases. The most frequent grade 3–4 toxicities observed were hematological, mainly neutropenia. Although cardiac toxicities are not frequently reported (20,32,33), six patients in our study experienced cardiac issues, all of whom had previously received anthracyclines that may increase susceptibility to cardiotoxicity. The European Medicines Agency recommends assessing left ventricular ejection fraction (LVEF) before treatment. Trabectedin should be withheld if LVEF declines by 15% or more (9). We recommend routine cardiac monitoring, including regular echocardiographic follow-ups, and scheduling a specialized cardiologist appointment at trabectedin initiation and upon any emergence of toxicity. Although observed in a single patient, it is also important to note that trabectedin is a vesicant, meaning it has the potential for extravasation from blood vessels, leading to subsequent damage in surrounding tissue (34,35). Extra vigilance is required during infusion.

Trabectedin was initially available through a compassionate use program starting in 2003 (18). However, in 2013, the French health authority removed trabectedin from the list of reimbursed drugs for advanced STS due to its limited medical benefit. A derogatory funding was granted until 2016 in France. Several studies have demonstrated that trabectedin is a cost-effective treatment option for advanced STS patients (16,36,37). Due to the results of ET743-SAR-3007 and T-SAR clinical trials and the absence of comparison to pazopanib and eribulin in the therapeutic arsenal, trabectedin was removed from the list of reimbursed drugs in France (12). The JCOG1802 study is a randomized trial designed to evaluate the effectiveness of trabectedin, eribulin, and pazopanib as second-line therapies for advanced STS (38). This phase II study was recently presented in a congress abstract, indicating that trabectedin demonstrated better PFS, DCR and overall survival compared to eribulin. However, it showed worse PFS, DCR and overall survival compared to pazopanib as the second-line treatment for patients with advanced STS (39). In several countries, the cost-effectiveness ratio has become a pressing concern in public health management, particularly for treatments lacking robust medical evidence (20,40). Other molecules have been withdrawn from reimbursement in France, notably cemiplimab for cutaneous squamous cell carcinoma (41). Unlike cemiplimab, which has alternative anti-PD-1 immunotherapies approved for this indication, trabectedin has no other options within the same therapeutic class. The high costs of treatment currently pose significant barriers to patient accessibility, especially considering the limited resources available within health systems and the increasing burden of cancer. Between 2014 and 2024, the retail price of trabectedin in France decreased substantially, from €1,600 to €180 per milligram (excluding taxes). Although it is no longer reimbursed, the gradual reduction in trabectedin prices facilitates its use in clinical practice, which is important given the benefits for patients with STS. In France, it is still possible to prescribe and order trabectedin, but the costs are covered by the hospital. Therefore, to control trabectedin costs related to STS treatments, the hospital management must approve each new treatment and a cost monitoring was implemented. Although the cost of trabectedin can be mitigated by the expenses related to hospitalization, the hospital will still incur a surcharge. The benefit-to-cost ratio is shifting, with increasingly low costs and a clinical benefit in terms of PFS of approximately 4.1 months, with relatively low toxicities. While these data might be methodologically insufficient to reapply for reimbursement, it would be worthwhile if this question could be addressed. Nevertheless, trabectedin clearly represents an additional line of treatment in the STS therapeutic arsenal, offering tangible real-life benefits.

This study is limited by its retrospective design, small sample size, and missing data, particularly concerning the association between histological subtypes and clinical outcomes. Furthermore, we focused on price per milligram of trabectedin vials. Costs related to hospitalization (€7,082 for average cost of stay in case of disease-related group #08M24), supportive treatments (e.g. dexamethasone, granisetron), and the management of trabectedin-induced side effects must also be considered. Despite these limitations, this study provides valuable insights for STS management, even though it is a very rare disease. Another important limitation is the heterogeneity of non-L-sarcomas, which are a disparate group of STS, with different histological subgroups potentially exhibiting varying responses to trabectedin. The limited statistical power of our analysis emphasizes the need for a multicenter study specifically focused on assessing the efficacy of trabectedin in non-L-sarcomas.

In conclusion, trabectedin provides a meaningful clinical benefit for STS patients with manageable toxicities. This study confirms the preferential use of trabectedin for L-sarcomas, especially myxoid liposarcomas. Still, trabectedin remains one of the few treatment options available for patients with non-L-sarcoma, extending progression-free survival by 2.4 months. A downward trend in the cost of this old drug is essential for its continued use and for improving the life expectancy of STS patients.

## Data Availability

All data produced in the present work are contained in the manuscript

## Additional information

## Acknowledgements

We would like to thank the participating patients.

## Authors’ contributions

JSG: Conceptualization, Data curation, Formal analysis, Investigation, Visualization, Writing – original draft, Writing – review and editing

SW: Conceptualization, Data curation, Formal analysis, Project administration, Supervision, Writing – original draft, Writing – review and editing

AA: Formal analysis, Writing – original draft, Writing – review and editing

VL: Data curation, Investigation, Writing – review and editing

DT: Data curation, Investigation, Writing – review and editing

SB: Data curation, Investigation, Writing – review and editing

SEZ: Data curation, Investigation, Writing – review and editing

NN: Data curation, Investigation, Writing – review and editing

CC: Conceptualization, Formal analysis, Writing – original draft, Writing – review and editing

RD: Conceptualization, Resources, Formal analysis, Project administration, Supervision, Writing – original draft, Writing – review and editing

CB: Conceptualization, Data curation, Formal analysis, Project administration, Supervision, Writing – original draft, Writing – review and editing

## Ethics approval and consent to participate

Eligible patients had signed a non-opposition consent form. The study was performed in accordance with the Declaration of Helsinki.

## Consent for publication

Not applicable

## Data availability

Not applicable

