## Supplemental Files for "Trabectedin in the Treatment of Soft Tissue Sarcoma: Real-world Data on Effectiveness, Safety, and Financial Implications from a European Comprehensive Cancer Centre"

**Supplementary Figure S1**. Market price (€ excluding taxes) per 1 mg bottle according to procurement period

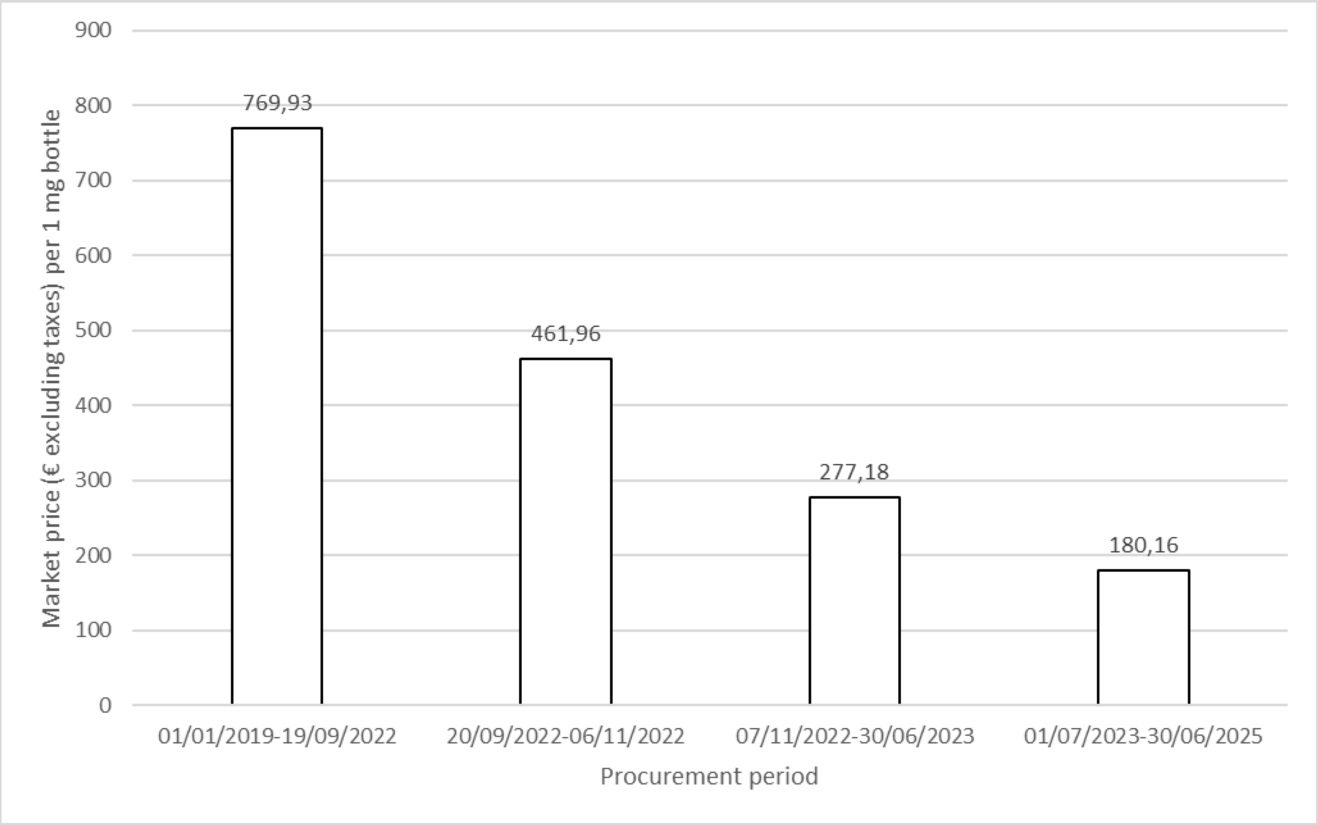

**Supplemental Figure S2**. Best responses to trabectedin according to the main classification of STS subtypes

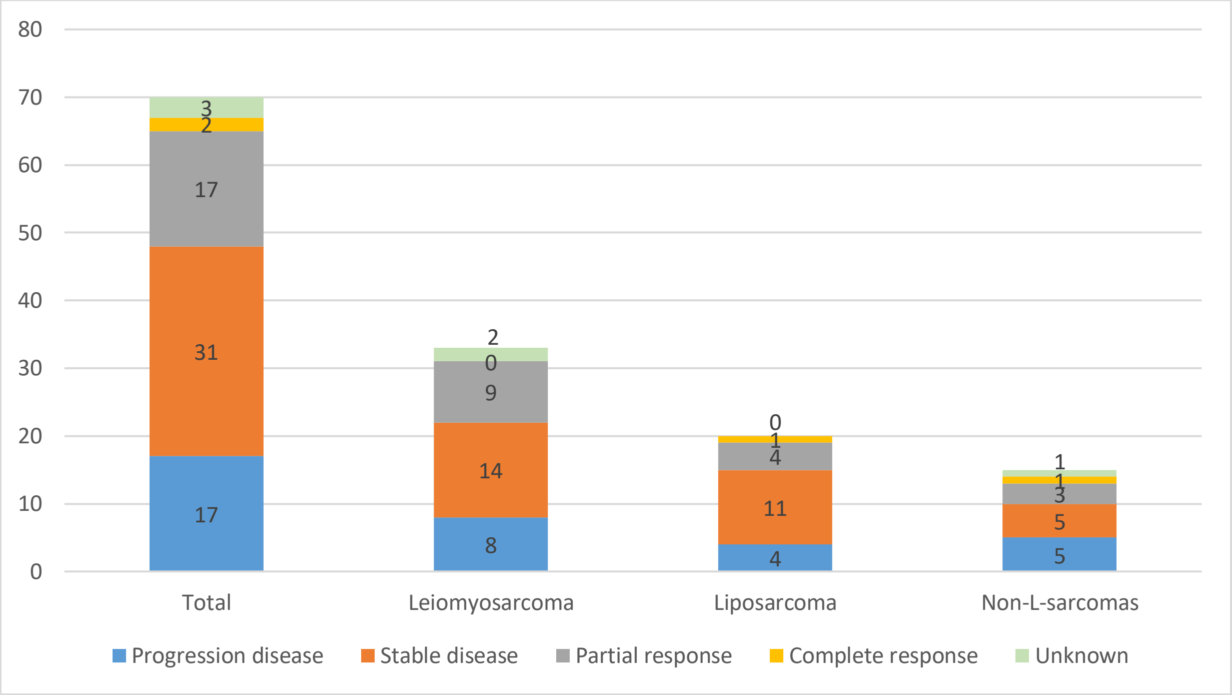

**Supplementary Figure S3. Progression-free survival (in days) according to the soft-tissue sarcoma subtypes**

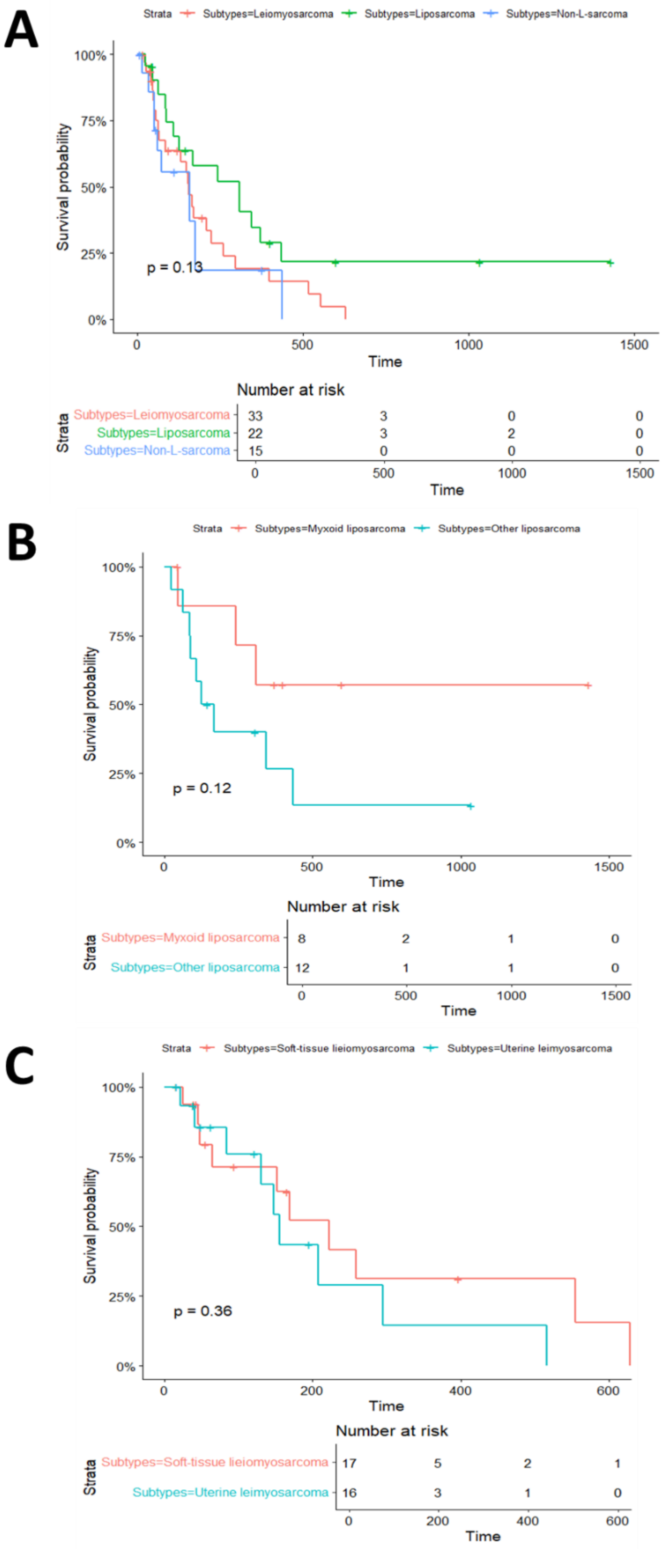


(A) Progression-free survival (in days) between leiomyosarcoma, liposarcoma and non-L-sarcoma. (B) Progression-free survival (in days) between myxoid liposarcoma and other liposarcoma. (C) Progression-free survival (in days) between uterine and soft-tissue leiomyosarcomas

**Supplementary Table S1. Trabectedin best response assessment in soft tissue sarcomas**

| N (%) | Leiomyosarcoma (n=33) | Liposarcoma (n=20) | Non-L-sarcoma (n=15) |
|---|---|---|---|
| Complete response | 0 (0) | 1 (5) | 1 (7) |
| Partial response | 9 (27) | 4 (20) | 3 (20) |
| Stable disease | 14 (43) | 11 (55) | 5 (33) |
| Progressive disease | 8 (24) | 4 (20) | 5 (33) |
| Unknown | 2 (6) | 0 (0) | 1 (7) |
| Objective response rate | 9 (27) | 5 (25) | 4 (27) |
| Disease control rate | 23 (70) | 16 (80) | 9 (60) |

**Supplementary Table S2. Compared survival between L-sarcoma subtypes**

| STS | Subtypes | Number of patients | PFS: median, months | HR [95%CI] | p-value |
|---|---|---|---|---|---|
| Leiomyosarcoma | Uterine | 17 | 3.1 | / | / |
| | Soft-tissue | 16 | 3.4 | 0.4459 [0.5981 - 4.079] | 0.363 |
| Liposarcoma | Myxoid | 8 | 11.3 | / | / |
| | Other | 12 | 4.5 | 1.01 [0.3633-0.7322] | 0.134 |

HR: hazard ratio; CI: confidence interval

**Supplementary Table S3. Cohort comparison of trabectedin use in advanced STS**

| Authors | PMID | Year of publication | Study | STS | Median PFS (months) | ORR (%) | DCR (%), evaluation |
|---|---|---|---|---|---|---|---|
| Samuels et al. | 23385197 | 2013 | Worldwide expanded access program | STS (n=807) | NA | 5.9 | 48.5, BR |
| | | | | L-sarcoma (n=476) | NA | 6.9 | 54.2, BR |
| | | | | Non-L-sarcoma (n=302) | NA | 4.0 | 37.7, BR |
| Blay et al. | 23388156 | 2013 | French compassionate use program | STS (n=181) : 104 L-sarcoma & 77 non-L-sarcoma | 3.6 | 10.0 | 49.0, BR |
| Hoiczyk et al. | 23652821 | 2013 | Retrospective, multicenter | STS (n=101) | 2.1 | NA | NA |
| | | | | L-sarcoma (n=46) | 3.1 | NA | NA |
| | | | | Non-L-sarcoma (n=55) | 1.6 | NA | NA |
| Le Cesne et al. | 25727882 | 2015 | Retrospective, multicenter | STS (n=804) | 4.4 | 16.5 | 66.7, BR |
| | | | | L-sarcoma (n=481) | 5.7 | 18.6 | 72.6, BR |
| Moriceau et al. | 26384694 | 2015 | Retrospective, monocenter | STS (n=59) | 2.7 | 6.8 | 23.7, BR |
| | | | | L-sarcoma (n=32) | 2.7 | NA | NA |
| | | | | Non-L-sarcoma (n=27) | 2.7 | NA | NA |
| Demetri G et al. | 26371143 | 2016 | **Phase III clinical trials** | L-sarcoma (n=345) | 4.2 | 9.9 | 61.2, BR |
| Buonadonna et al. | 28926423 | 2017 | Prospective, multicenter | STS (n=218) : 143 L-sarcoma & 75 non-L-sarcomas | 5.9 | 26.6 | 65.6, BR |
| Shamai et al. | 30324774 | 2018 | Retrospective, monocenter | STS (n=86) : L-sarcoma (n=77) & Non-L-sarcoma (n=9) | NA | 22.1 | 54.7, BR |
| Kobayashi et al. | 31825533 | 2020 | Retrospective, multicenter | STS (n=140) | 3.7 | NA | NA |
| | | | | L-sarcoma (n=60) | 8.4 | NA | NA |
| | | | | Non-L-sarcoma (n=80) | 3.2 | NA | NA |
| Palmerini et al. | 33801399 | 2021 | Retrospective, multicenter | STS (n=512) | 5.1 | 13.7 | 46.7, BR |
| | | | | L-sarcoma (n=348) | 8.3 | 16.1 | 53.4, BR |
| | | | | Non-L-sarcoma (n=164) | 2.4 | 8.5 | 32.3, BR |
| Le Cesne et al. | 33932507 | 2021 | **Phase III clinical trials** | STS (n=52) | 3.1 | 13.7 | 80.4, BR |

| | | | | | | | |
|---|---|---|---|---|---|---|---|
| | | | | L-sarcoma (n=32) | 5.1 | 21.9 | 87.5, BR |
| | | | | Non-L-sarcoma (n=20) | 1.8 | NA | 64.8, BR |
| Chaigneau et al. | 36283345 | 2022 | Retrospective, multicenter | STS (n=126) | 3.0 | 9.7 | 46.0, BR |
| | | | | L-sarcoma (n=75) | 2.8/2.3 | NA | NA |
| | | | | Non-L-sarcoma (n=51) | 1.7 to 13.3 | NA | NA |
| Cerdà Serdà et al. | 35129790 | 2022 | Retrospective, monocenter | Non-L-sarcoma (n=34) | 2.9 | 8.8 | 32.4, BR? |
| Grünwald et al. | 36358652 | 2022 | Prospective, multicenter | STS (n=128) : 68 L-sarcoma & 60 non-L-sarcoma | 5.2 | 11.7 | 45.3, BR |
| Ön et al. | 39590133 | 2024 | Retrospective, multicenter | L-sarcoma (n=98) | 3.0 | 16.0 | 42.0, unknown |

PMID: Pubmed identifier. STS: Soft Tissue Sarcoma. PFS: Progression-Free Survival. ORR: Objective Response Rate. DCR: Disease Control Rate. BR : best response
